# Population Changes in Seroprevalence among a Statewide Sample in the United States

**DOI:** 10.1101/2020.12.18.20248479

**Authors:** Kristen Malecki, Maria Nikodemova, Amy Schultz, Matt Walsh, Andrew J. Bersch, Ajay K. Sethi, Paul E. Peppard, Corinne Engelman, Lisa Cadmus-Bertram, Nasia Safdar, Allen Batemen, Ryan Westergaard

## Abstract

Antibody surveillance provides essential information for public health officials to work with communities to discuss the spread and impact of COVID-19. At the start of the new severe acute respiratory syndrome coronavirus 2 (SARS-CoV-2) pandemic in the United States, diagnostic testing was limited with many asymptomatic and thus undetected cases. Irrespective of symptom severity, antibodies develop within two to three weeks after exposure and may persist 6 months or more.; Thus, antibody surveillance is an important tool for tracking trends in past infections across diverse populations. This study includes adults and children (≥12 years old) recruited from a statewide sample of past 2014-2020 Survey of the Health of Wisconsin (SHOW) participants. SHOW, an ongoing population-based health examination study including a randomly selected sample of households, partnered with the Wisconsin Department of Health Services and the Wisconsin State Laboratory of Hygiene to conduct longitudinal antibody surveillance using the Abbott Architect SARS-CoV-2 IgG antibody test, which detects antibodies against the nucleocapsid protein. Three WAVES of sample collection were completed in 2020-2021, tracking mid-summer, late fall, and early spring COVID-19 trends prior to vaccine availability. Crude estimates of seroprevalence in the total study population increased ten-fold from 1.4% during WAVE I to 11.5% in WAVE III. Within the statewide probability sample, weighted estimates increased from 1.6% (95% CI:0.6-2.5%), to 6.8% (95% CI:4.3-9.4%) in WAVE II and to 11.4% (95% CI:8.2, 14.6%) in WAVE III. Longitudinal trends in seroprevalence match statewide case counts. Local seroprevalence showed variation by state health region with increasing prevalence among higher income (>200% poverty income ratio), and rural health regions of the state seeing the highest increase in COVID-19 prevalence over time. Significant disparities in prevalence by racial and ethnic groups also exist, with greater than two times seroprevalence among Latino and black participants compared to non-Hispanic whites. This public health and academic partnership provides critical data for the ongoing pandemic response and lays the foundation for future research into longer-term immunity, health impacts and population-level disparities.

## Background

In the United States, the COVID-19 pandemic caused by the new severe acute respiratory syndrome coronavirus 2 (SARS-CoV-2) was recognized as a significant public health threat in March of 2020. Scaling up of diagnostic testing was slow at the start, particularly in non-urban settings and non-hospitalized individuals. Community spread is linked to many asymptomatic or mild cases [1]. Consequently, case counts based on antigen or PCR testing alone likely underestimate the true number of infections. Antibodies develop within two to three weeks after the exposure [2] for SARS-CoV-2 and may persist 6 months or more in some individuals [3-5], however, a decline was observed in some individuals within 3-5 months after infection [6-10]. By providing an indication of past infection, antibody surveillance therefore provides a means to examine trends in COVID-19 spread in the population over time [11]. Very few studies have state-wide data, aggregated by local health region, characterized by demographic factors and gathered longitudinally to support population health monitoring and impacts of COVID-19. Local data can support a more targeted public health response. The present study, which consists of local state-wide data, contributes to our understanding of the use of antibody surveillance and the persistence of antibodies over time.

## Methods

This study includes a longitudinal sample of adults and children (≥12 years old on June 21, 2021) recruited from the statewide Survey of the Health of Wisconsin (SHOW) study. Individuals were eligible if they had participated in SHOW health examination survey between 2014-2016 or were included in the special population oversample conducted in the city of Milwaukee during 2018-2019. This antibody surveillance study was designed in partnership with the Wisconsin Department of Health Services (DHS).

Three waves of repeated blood sample collection among SHOW participants were conducted across the entire state. WAVE I collection occurred in early July through August 2020; WAVE II began in late October and continued through early December; WAVE III was conducted March-April 2021. Twelve sample collection sites were identified across Wisconsin (e.g., health departments, churches, and community-based organizations) and both urban and rural locations were included. Data collection was conducted in English except for one site in Milwaukee that offered Spanish interpreters. Each site was equipped with a small cooler and centrifuge. Participants were screened for fever, COVID-19 symptoms and known past exposures. Those with active COVID-19 infection or COVID-19 exposure within the past 2 weeks were excluded. Participants completed a short questionnaire followed by venous blood collection by trained phlebotomists. Blood was processed to serum in the field and kept at 4°C. Samples were collected by courier each evening and sent immediately to the Wisconsin State Laboratory of Hygiene, where they were analyzed using the Abbott Architect SARS-CoV-2 IgG antibody test, which has an estimated specificity of 99.6% [11, 12]. Participants were consented for use of questionnaire data and additional blood sample collection for future COVID-19 research. The study was approved by the University of Wisconsin Health Sciences Institutional Review Board (IRB). Further details on study protocols are available from study investigators.

## Findings

A total of 1056, 1070, and 1002 individuals were tested in WAVE I through WAVE III respectively, corresponding to an average response rate of 39-40% among all eligible participants. Figures 1 shows the overlap of participants across all three WAVES. Of these, 996, 994, and 930 participants were in the original statewide population-based probability sample and were used to estimate weighted statewide estimates reported in Table 1. From WAVE I to WAVE II, crude estimates of seroprevalence in the total study population increased from 1.9% to 7.3%. Within the statewide probability sample, weighted estimates increased from an estimated 1.6% (95% CI:0.6-2.5%) seropositivity to 6.8% (95% CI:4.3-9.4%). These data were collected during late October and early November, a time in which there was a significant increase in overall documented cases in the state. By March/April 2021, seroprevalence had doubled to 11.4% (95% CI:8.2-14.6%)

**Table 1.** Past Antibody COVID-19 Community Survey Seroprevalence Results, WAVE I, WAVE II and WAVE III

|  | WAVE I July -Early August, 2020 |  |  |  | WAVE II Late October- Early December, 2020 |  |  |  | WAVE III Late March- April, 2021 |  |  |  |
| --- | --- | --- | --- | --- | --- | --- | --- | --- | --- | --- | --- | --- |
| Select Factors | N | Crude Percent | Weighted Percent* | Weighted 95% CI* | N | Crude Percent | Weighted Percent* | Weighted 95% CI* | N | Crude Percent | Weighted Percent* | Weighted 95% CI* |
| TOTAL | 996 | 1.4 | 1.6 | (0.6, 2.5) | 994 | 6.5 | 6.8 | (4.3,9.4) | 929 | 11.5 | 11.4 | (8.2, 14.6) |
| Age (on June 21, 2020) |  |  |  |  |  |  |  |  |  |  |  |  |
| 12 to 44 | 309 | 1.3 | 1.4 | (0.0, 2.9) | 297 | 5.7 | 6.1 | (1.5, 10.8) | 261 | 10.7 | 10.5 | (5.0, 16.0) |
| 45 to 64 | 308 | 2.0 | 1.9 | (0.2, 3.6) | 318 | 8.2 | 8.1 | (5.6, 10.7) | 301 | 13.3 | 12.9 | (7.6, 18.2) |
| 65 or older | 379 | 1.1 | 1.5 | (0.1, 3.0) | 379 | 5.8 | 6.4 | (3.3, 9.5) | 367 | 10.6 | 11.2 | (8.1, 14.3) |
| Gender |  |  |  |  |  |  |  |  |  |  |  |  |
| Male | 418 | 1.2 | 1.6 | (0.8, 3.2) | 405 | 8.2 | 9.4 | (4.3, 14.5) | 387 | 11.9 | 11.5 | (6.9, 16.1) |
| Female | 578 | 1.7 | 1.6 | (0.8, 3.2) | 589 | 5.4 | 4.3 | (2.7, 5.8) | 542 | 11.3 | 11.3 | (7.0, 15.6) |
| Race / Ethnicity |  |  |  |  |  |  |  |  |  |  |  |  |
| Non-Hispanic white (alone) | 870 | 1.2 | 1.3 | (0.1, 2.4) | 861 | 6.3 | 6.5 | (3.8, 9.2) | 817 | 11.5 | 10.8 | (7.9, 13.6) |
| Non-Hispanic Black or African American (alone or in combination) | 50 | 4.0 | 5.6 | (0.0, 14.5) | 65 | 7.7 | 4.1 | (0.0, 8.5) | 45 | 8.9 | 18.7 | (0.5, 36.9) |
| Hispanic (any race) | 20 | 5.0 | 2.3 | (0.0, 7.0) | 23 | 8.7 | 12.8 | (0.0, 32.1) | 22 | 13.6 | 11.5 | (0.0, 23.7) |
| Non-Hispanic other or multiracial (not Black or African American) | 56 | 1.8 | 1.0 | (0.0, 3.1) | 45 | 8.9 | 7.8 | (0.0, 20.0) | 45 | 13.3 | 12.7 | (0.0, 27.2) |
| Poverty (at previous survey 2014-2019) |  |  |  |  |  |  |  |  |  |  |  |  |
| ≥200% FPL | 780 | 1.0 | 1.1 | (0.1, 2.0) | 768 | 6.8 | 7.5 | (4.2, 10.7) | 728 | 12.6 | 13.2 | (10.2, 16.3) |
| <200% FPL | 216 | 2.8 | 2.7 | (0.3, 5.1) | 226 | 5.8 | 5.2 | (1.1, 9.4) | 201 | 7.5 | 6.9 | (1.7, 12.2) |

| Health Region |  |  |  |  |  |  |  |  |  |  |  |  |
| --- | --- | --- | --- | --- | --- | --- | --- | --- | --- | --- | --- | --- |
| SE | 341 | 2.8 | 2.7 | (0.3, 5.1) | 346 | 7.8 | 9.2 | (3.3, 15.1) | 305 | 11.5 | 10.8 | (3.7, 17.8) |
| S | 94 | 2.6 | 3.1 | (1.3, 4.9) | 98 | 4.1 | 2.4 | (0.6, 4.3) | 96 | 8.3 | 9.4 | (1.3, 17.4) |
| W | 288 | 0.0 | 0.0 | NA | 276 | 4.0 | 3.9 | (2.9, 4.9) | 258 | 10.9 | 11.6 | (5.5, 17.7) |
| N & NE | 273 | 0.3 | 0.3 | (0.0, 0.7) | 274 | 8.4 | 8.7 | (5.0, 12.4) | 270 | 13.3 | 13.0 | (8.6, 17.4) |
| * Estimates accounted for the sampling weights and sampling design elements of stratification and clustering. |  |  |  |  |  |  |  |  |  |  |  |  |

**Table 2.** Surveillance Recruitment and Response Rates for COVID-19 Antibody Testing*

| Sample WAVE I | Eligible | Participate | Response Rate (%) | Positive Tests | % Positive |
| --- | --- | --- | --- | --- | --- |
| 2014-2016 | 2228 | 952 | 42.7 | 13 | 1.4 |
| 2018-2019 (probability) | 273 | 47 | 17.2 | 1 | 2.1 |
| 2018-2019 (convenience) | 169 | 33 | 19.5 | 2 | 6.1 |
| Latinx pilot | 104 | 24 | 23.1 | 4 | 16.7 |
| Sample WAVE II | Eligible | Participate | Response Rate | Positive Tests | % Positive |
| 2014-2016 | 2228 | 930 | 41.7 | 61 | 6.6 |
| 2018-2019 (probability) | 273 | 64 | 23.4 | 4 | 6.3 |
| 2018-2019 (convenience) | 169 | 38 | 22.5 | 3 | 7.9 |
| Latinx pilot | 104 | 38 | 36.5 | 10 | 26.3 |
| Sample WAVE III** | Eligible | Participate | Response Rate | Positive Tests | % Positive |
| 2014-2016 | 2092 | 883 | 42.2 | 103 | 11.7 |
| 2018-2019 (probability) | 220 | 47 | 21.4 | 4 | 8.5 |
| 2018-2019 (convenience) | 112 | 40 | 35.7 | 8 | 20.0 |
| Latinx pilot | 71 | 32 | 45.1 | 3 | 9.4 |

\*NOTE: Study sample individuals were asked to refrain from participation if they had an active COVID-19 infection or known COVID-19 exposure in the last two weeks.
\*\*The eligible participants are lower than waves 1 & 2 due to ongoing cohort maintenance (ie. Removing deceased individuals, etc).

**Figure 1.**
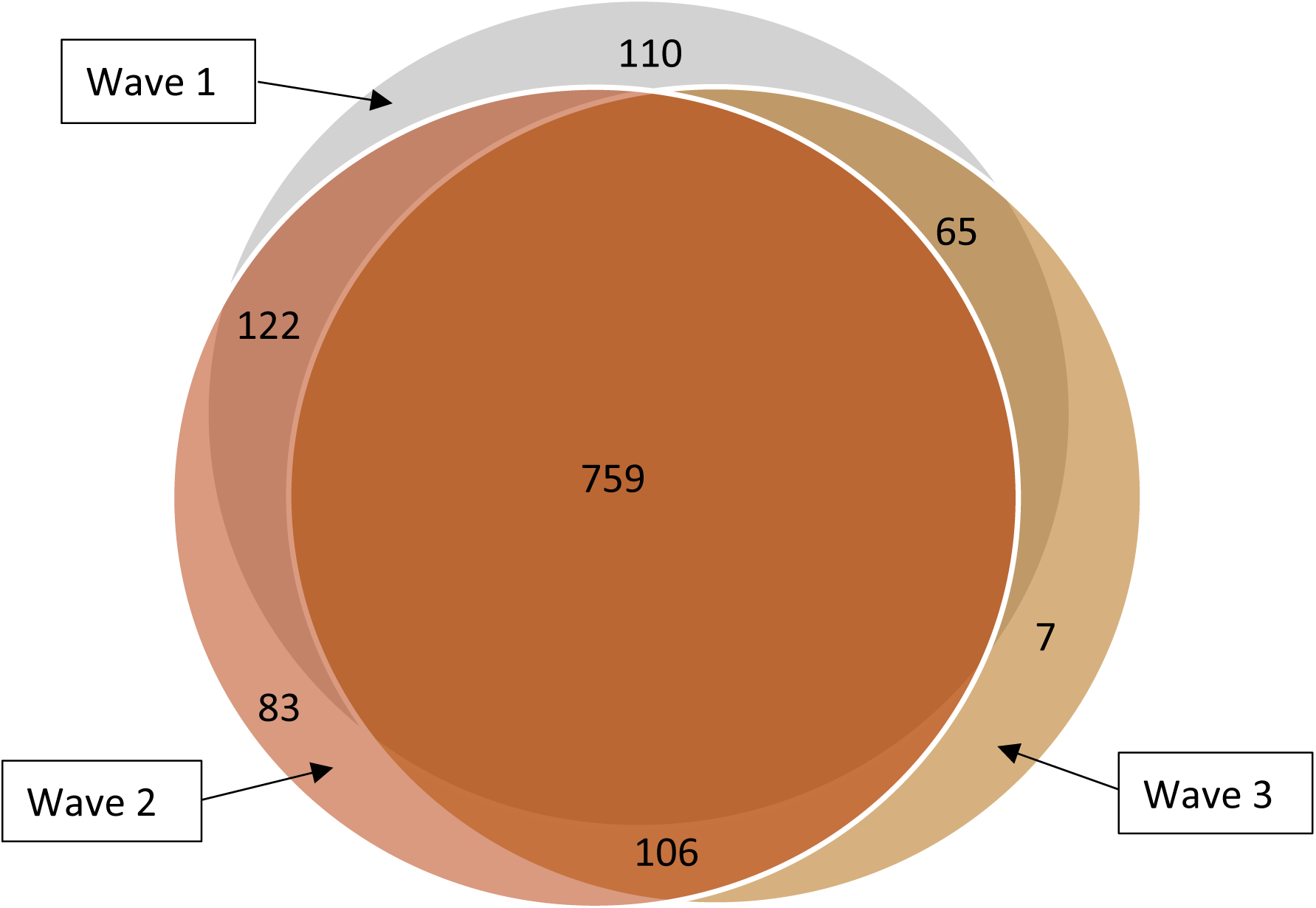
Overlap among participants from PACCS Waves I, II and III.

Seroprevalence varied by age and gender across all three WAVES with increasing prevalence among children in WAVE III. Although there was no gender difference observed in WAVE I, seropositivity rates was higher in men 9.4% (95% CI:4.3, 14.3) compared to women 4.3% (2.7-5.3%) in WAVE II. The highest among 45-64 year-olds at 8.1% (5.6-10.7%) up from 1.9% positive in WAVE I and compared to 6.1% (1.5-10.8%) among participants younger than 44 and 6.4% positive (3.3-9.5%) in those older than 65. There were 59 participants 12-17 years of age in WAVE I, 65 in WAVE II and 57 in WAVE III. No children were positive in the WAVE I, the seroprevalence among child participants in WAVE II was 9.2%. Among WAVE III, the seroprevalence among child participants increased to 19.3%.

Sub-population differences in seroprevalence were observed by state agency designated health regions and by racial/ethnic composition and location of study participants. Health regions are defined by the Wisconsin Department of Health Services to represent geographic service areas for local and state public health practitioners. In WAVE I, the greatest proportion of positive tests was detected in the Southeastern region of the state, home to Milwaukee, the largest metropolitan area in the state (3.1% positive; 95% CI:1.4-4.9%); this increased to 9.3% (95% CI:3.3-15.1%) in WAVE II and was 10.8 (95% CI: 3.7-17.8%) in WAVE III. The greatest change in antibody-positive results from WAVE I to WAVE II was in the North and Northeast from 1.7% (0.0-4.3%) to 8.7% (95% CI: 5.0-12.4%), this jump persisted through WAVE III to 13.0% (8.6, 17.4%).

Difference in prevalence also varied by self-identified race-ethnicity and income across three waves. Non-whites (n=133) also had a higher prevalence of 8.3% (0.3-16.4%) compared to Non-Hispanic Whites (n=861) with 6.5% (3.8-9.2%). Among the non-white participants, n=55 were among a Latino population oversample recruited from a largely industrial and working-class community on Milwaukee’s south side. Of these individuals, over 25% had positive antibodies measured during WAVE II, by WAVE III seroprevalence was reduced to 9%. Among other Milwaukee residents who largely self-identified as black or African American we saw shifts in seroprevalence from 6.1% in WAVE 1 to 20.0% in WAVE III. Seroprevalence by income above and below 200% of the federal poverty level (FPL) appeared to higher in lower income individuals (living below 200% FPL) but during WAVE III these trends, while not statistically different appeared to reverse with greater percent seroprevalence in higher income individuals.

A total of 876 individuals participated in both WAVE I and WAVE II and 759 in all three WAVES. Figure 2 demonstrates positivity results by WAVE of survey participation, including those who did not complete a second WAVE (missing), positive or negative. Of the 18 that tested positive for antibodies in WAVE I and participated in WAVE II, 6 (33%) had a negative antibody test in WAVE II. Among the 7 that remained positive in WAVE II and complete WAVE III 4 were positive, and 3 (43%) had a negative antibody test.

**Figure 2.**
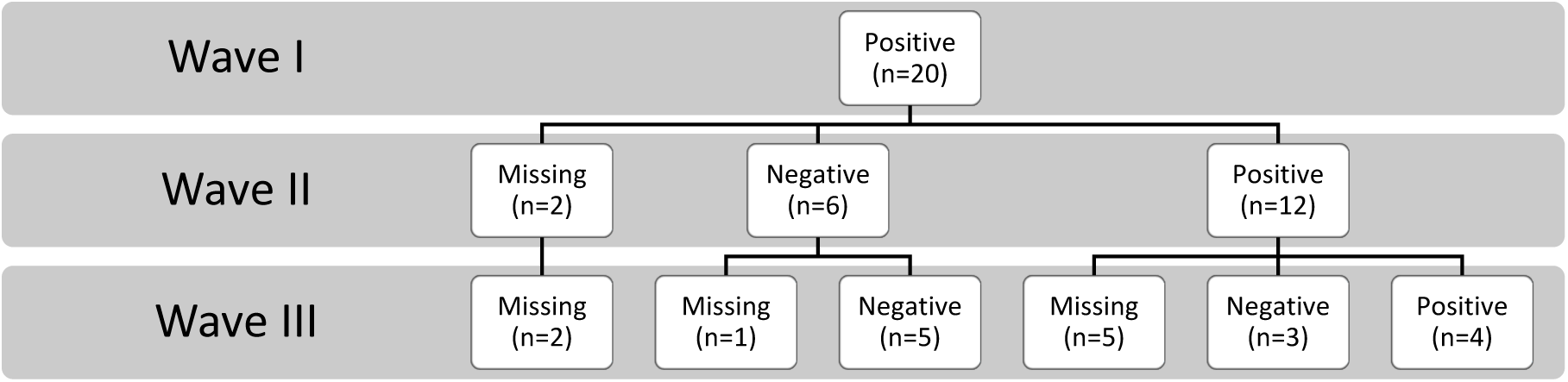
Positivity Among WAVE I Positives.

## Interpretation

Results show an overall four to fivefold increase in the prevalence of COVID-19 antibodies across the state of Wisconsin from the first sample collection in July and August to the second sample collection in later October through early December 2020. Regional variation in trends is consistent with statewide testing results and mirror the overall trends in COVID-19 infections in Wisconsin. The antibody testing suggests that as of early November 2020, just prior to a second statewide spike in COVID-19 infections, over 90% of the state’s population did not have SARS-CoV-2 antibodies. This may be due to lack of exposure to SARS-CoV-2 or because infection had occurred, but antibodies were no longer detectable. The disparities in seroprevalence among special population samples indicate significantly higher burden in these groups, likely resulting from higher representation of front-line essential workers, health care employees, and households living in more crowded conditions. However, additional research is needed to better understand the significant sources of disparities among these populations. While it is unclear how long individuals will have detectable levels of antibodies after an infection of COVID-19, results do reflect the increasing trends of COVID-19 infections that occurred between late Spring and Fall across the state of Wisconsin. Further investigation regarding symptom severity, exposure and lingering side effects will provide additional insights into the long-term impacts of COVID-19 infection on immunity and health outcomes.

Three WAVEs of sample collection among a representative sample of state residents offered a unique opportunity to track sub-population changes. In particular, changes by state health agency designated regions and changes in child prevalence over time suggest that risk of infection across time varied across region and demographic populations in states. These sub-population analyses are important for public health practitioners in targeting outreach and in understand risks over time. Further analyses of questionnaire data on vaccine hesitancy and symptoms among those adults and children to determine symptom severity and awareness of past infections even if asymptomatic will offer additional insights into how the pandemic unfolded within a state population. These data are also important in considering emergence of new variants and longevity of natural immunity from infections overtime.

## Data Availability

All data are available from study investigators upon request.

https://show.wisc.edu/

## Research in context

This is one of a few local and state-wide population-based studies generated from a random statewide representative sample of adults and children to show that antibody prevalence follows infections in the population. Our observation that one third of participants who tested antibody positive at WAVE I did not retest positive at WAVE II suggests that antibodies are not always long-lasting, and the implications for long-term immunity remain unknown. It is also important to point out that some changes could be due to specificity of the test itself. At 99.6% specificity, we would expect a portion of the tests in both rounds are due to false positives, but it is hard to ascertain how this is truly impacting study results [13, 14]. Nonetheless, these data provide essential information for public health officials to work with local communities to discuss the spread and impact of COVID-19. Moving forward, the long-term implications of past infections on health and well-being can be monitored as part of the ongoing Survey of the Health of Wisconsin research program.

## Acknowledgements

We would like to acknowledge all past SHOW participants willing to take place in this surveillance effort. Additional acknowledgements to SHOW Program Manager Jen Tratnyek and her team for supporting logistics, the SHOW phlebotomy coordinator Jonathan Cabezas-Olcoz, and field coordinator Doug Esselman, and lead administrative support from Ben Young for study organization, recruitment and coordination, Allison Rodriguez for Spanish translation and Jacqueline Cronin for data dissemination and outreach efforts. Further thanks to the various sample collection site partners across the state of Wisconsin who opened their doors for this work. The Wisconsin State Laboratory of Hygiene staff also played a key role in supporting these efforts in coordination with the SHOW program and were responsible for coordination of testing and reporting of results to the SHOW program for further dissemination of results to participants.

## REFERENCES

1. Long, Q.-X., et al., Clinical and immunological assessment of asymptomatic SARS-CoV-2 infections. Nature Medicine, 2020. 26(8): p. 1200–1204.

2. Wu, J., et al., SARS-CoV-2 infection induces sustained humoral immune responses in convalescent patients following symptomatic COVID-19. medRxiv, 2020: p. 2020.07.21.20159178.

3. Dan, J.M., et al., Immunological memory to SARS-CoV-2 assessed for up to eight months after infection. bioRxiv, 2020: p. 2020.11.15.383323.

4. Pradenas, E., et al., Stable neutralizing antibody levels six months after mild and severe COVID-19 episode. bioRxiv, 2020: p. 2020.11.22.389056.

5. Wajnberg, A., et al., Robust neutralizing antibodies to SARS-CoV-2 infection persist for months. Science, 2020. 370(6521): p. 1227–1230.

6. Ibarrondo, F.J., et al., Rapid Decay of Anti–SARS-CoV-2 Antibodies in Persons with Mild Covid-19. New England Journal of Medicine, 2020. 383(11): p. 1085–1087.

7. Korte, W., et al., SARS-CoV-2 IgG and IgA antibody response is gender dependent; and IgG antibodies rapidly decline early on. The Journal of infection, 2020: p. S0163-4453(20)30567-3.

8. Röltgen, K., et al., SARS-CoV-2 Antibody Responses Correlate with Resolution of RNAemia But Are Short-Lived in Patients with Mild Illness. medRxiv, 2020: p. 2020.08.15.20175794.

9. Seow, J., et al., Longitudinal evaluation and decline of antibody responses in SARS-CoV-2 infection. medRxiv, 2020: p. 2020.07.09.20148429.

10. Yin, S., et al., Longitudinal anti-SARS-CoV-2 antibody profile and neutralization activity of a COVID-19 patient. The Journal of infection, 2020. 81(3): p. e31–e32.

11. Ainsworth, M., et al., Performance characteristics of five immunoassays for SARS-CoV-2: a head-to-head benchmark comparison. The Lancet Infectious Diseases, 2020. 20(12): p. 1390–1400.

12. Bryan, A., et al., Performance Characteristics of the Abbott Architect SARS-CoV-2 IgG Assay and Seroprevalence in Boise, Idaho. Journal of Clinical Microbiology, 2020. 58(8): p. e00941–20.

13. Luchsinger, L.L., et al., Serological Assays Estimate Highly Variable SARS-CoV-2 Neutralizing Antibody Activity in Recovered COVID-19 Patients. Journal of Clinical Microbiology, 2020. 58(12): p. e02005–20.

14. McAdam, A., et al. SARS-CoV-2 Testing: Sensitivity Is not the Whole Story. 2020. 2020.

